# Sleep alterations as a function of 88 health indicators

**DOI:** 10.1101/2023.11.20.23298781

**Authors:** Péter Przemyslaw Ujma, Róbert Bódizs

## Abstract

Alterations in sleep have been described in multiple health conditions and as a function of several medication effects. However, evidence generally stems from small univariate studies. Here, we apply a large-sample, data-driven approach to investigate patterns between changes in sleep macrostructure, quantitative sleep EEG and health. We use data from the MrOS Sleep Study, containing polysomnography and health data from a large sample (N=3086) of elderly American men to establish associations between sleep macrostructure, the spectral composition of the electroencephalogram, 38 medical disorders, 2 health behaviors and the use of 48 medications. Of sleep macrostructure variables, increased REM latency and reduced REM duration was the most common finding across health indicators, along with increased sleep latency and reduced sleep efficiency. We found that the majority of health indicators were not associated with objective EEG PSD alterations. Associations with the rest were highly stereotypical, with two principal components accounting for 85-95% of the PSD-health association. PC1 consists of a decrease of slow and an increase of fast PSD components, mainly in NREM. This pattern was most strongly associated with depression/SSRI medication use and age-related disorders. PC2 consists of changes in mid-frequency activity. Increased mid-frequency activity was associated with benzodiazepine use, while decreases are associated with cardiovascular problems and associated medications, in line with immune-mediated circadian demodulation in these disorders. Specific increases in sleep spindle frequency activity were associated with taking benzodiazepines and zolpidem. Sensitivity analyses supported the presence of both disorder and medication effects.

## Introduction

Sleep problems are increasingly recognized as frequent ailments with a potentially great negative effect on both quality of life and economic productivity [1]. As sleep problems frequently appear later in life, the growth of the geriatric population makes sleep problems a significant, multifaceted public health challenge [2]. A further factor underscoring the importance of sleep problems is that they can be consequences or complications of pre-existing problems of physical or mental health, and may also arise as a side effect of medications.

Sleep problems can consist of reductions in either subjective or objective health quality. Subjective sleep quality is associated with wellbeing in older adults with multimorbidity, even after controlling the effects of other health-related factors [3]. Likewise, sleep quality was found to be an independent predictor of quality of life [4,5] or self-rated health [6], suggesting widespread associations between self-rated sleep and health.

Studies investigating changes in objective sleep indicators as a function of health or medication are scarce. Direct, disease-specific pathophysiological processes are known to affect sleep regulation in certain medical conditions. For example, Parkinson’s disease, epilepsy and other neuropsychiatric disorders are tightly linked with fundamental sleep regulatory mechanisms and are known to associate with specific alterations of EEG-based, objective sleep measures, sometimes even predicting disease-specific behavioral correlates [7–9]. Furthermore, some disorders affecting organs other than the brain (for example, the liver, the kidney or the heart) are known to cause changes in objectively rated sleep through both direct (physiological, e.g. altered melatonin synthesis) and indirect (symptom-mediated, e.g. pain affecting sleep) routes [10–12]. Furthermore, some pharmacological agents adopted in the treatment of various medical conditions are potent modulators of sleep (see [13,14] for reviews of the pharmaco-electroencephalographic literature on the effects of drugs applied in the treatment of neuropsychiatric conditions such as benzodiazepine hypnotics and SSRI antidepressants). Specific cases of drug-induced sleep EEG effects are benzodiazepines and benzodiazepine receptor agonists, also known as positive allosteric modulators of GABA_A_ receptors, which are known for their suppressive effects on low- and facilitating effects on high-frequency EEG activities in all behavioral states, with the additional increase of spindle frequency activity in NREM sleep [15–17]. A further widely reported medication effect on sleep is the REM-suppressive effect of several classes of antidepressive pharmacological agents, including selective serotonin reuptake inhibitors (SSRI), selective noradrenalin reuptake inhibitors (SNRI), tryciclic antidepressants (TCA) and monoamine oxidase inhibitor (MAOI) drugs [18,19]. In addition, several non-psychotropic medications were reported to cause sleep disturbances [20,21].

These studies, however, tend to be small and focus on only a specific health indicator or medication. Moreover, the data is typically derived from either non-medical settings (experiments) or clinical studies on highly selected samples. To our knowledge no large-scale, multivariate, ecologically valid (non-experimental) investigation on the associations of health conditions, medical treatments and sleep EEG is available in the literature. Many health problems potentially affecting sleep are under-researched, and studies on medication effects need replication on non-selected samples of patients, also taking into account multimorbidity and pharmacological polytherapy. Here, we leverage a large sample (N=3086) of elderly American men to investigate the associations between objectively measured sleep and a large set of health problems and medications. Given, the ubiquity of the spectral analysis of sleep-EEG [22], in addition to studying sleep macrostructure we also focus our work on the frequency composition of sleep EEG signals.

## Methods

### Participants and electrophysiology

We used data from the MrOS Sleep Study, downloaded from NSRR (www.sleepdata.org). The MrOS Sleep Study, an ancillary study of the main Osteoporotic Fractures in Men Study (MrOS) [23,24], is an investigation of 3135 elderly American men (mean age in current sample: 73.06 years, SD=5.55 years, N=3086 after exclusion of participants using CPAP, BiPAP or mouthpieces, or having undergone tracheoctomy or oxygen therapy in the past 3 months, as recommended for this cohort). Participants underwent full unattended polysomnography monitoring between December 2003 and March 2005. In these sessions, EEG was recorded from the central recording locations C3 and C4 using gold cup electrodes, originally recorded with an Fpz reference, re-referenced to a contralateral mastoid reference. A sampling frequency of 256 Hz and a high-pass hardware filter was used. All recordings were visually scored by experts (see [25] for further recording details). Artifacts were automatically rejected on a 4-second basis, based on extreme Hjorth parameters deviating by at least 2 standard deviations from the vigilance state average [26].

Sleep macrostructure was determined based on visual scoring by experts using standard criteria [27]. In our analyses, we used the following sleep macrostructure variables: clock time at sleep onset (expressed relative to midnight), total sleep time (TST), wake after sleep onset (WASO), sleep efficiency, sleep latency, REM latency, REM latency excluding wakefulness, and N1, N2, SWS and REM duration/percentage.

In each participant, we calculated power spectral density (PSD) as the main quantitative EEG outcome of interest. PSD calculation was implemented with the periodogram() function in MATLAB EEGLab [28], using 4 second epochs with 50% overlap and Hamming windows. PSD was calculated separately for NREM and REM and averaged across all epochs. PSD estimates were log10 transformed to normalize variances and relativized by subtracting the mean of all PSD values from each bin estimate in order to neutralize the effects of between-individual voltage differences.

### Health indicators

During the sleep visit, participants filled out a questionnaire in which they were asked about whether or not they have been diagnosed with 40 medical conditions. Data from four further conditions (arthritis/gout, cancer, surgical removal of stomach/intestines, and kidney stones) was available from the baseline MrOS visit and added to the list of conditions.

As part of the sleep study participants were invited to bring the medications they regularly take to a personal medical visit. The medications they presented were reviewed by the attending medical professional, who marked 48 common medications as present in or absent from the participants’ medication regimen.

Furthermore, participants reported the amount of coffee (in cups) and the number of cigarettes they consumed in the 4 hours preceding PSG recordings.

The full list of medical conditions and medications (henceforth jointly referred to as health indicators) is available in **Table 1**.

**Table 1.**
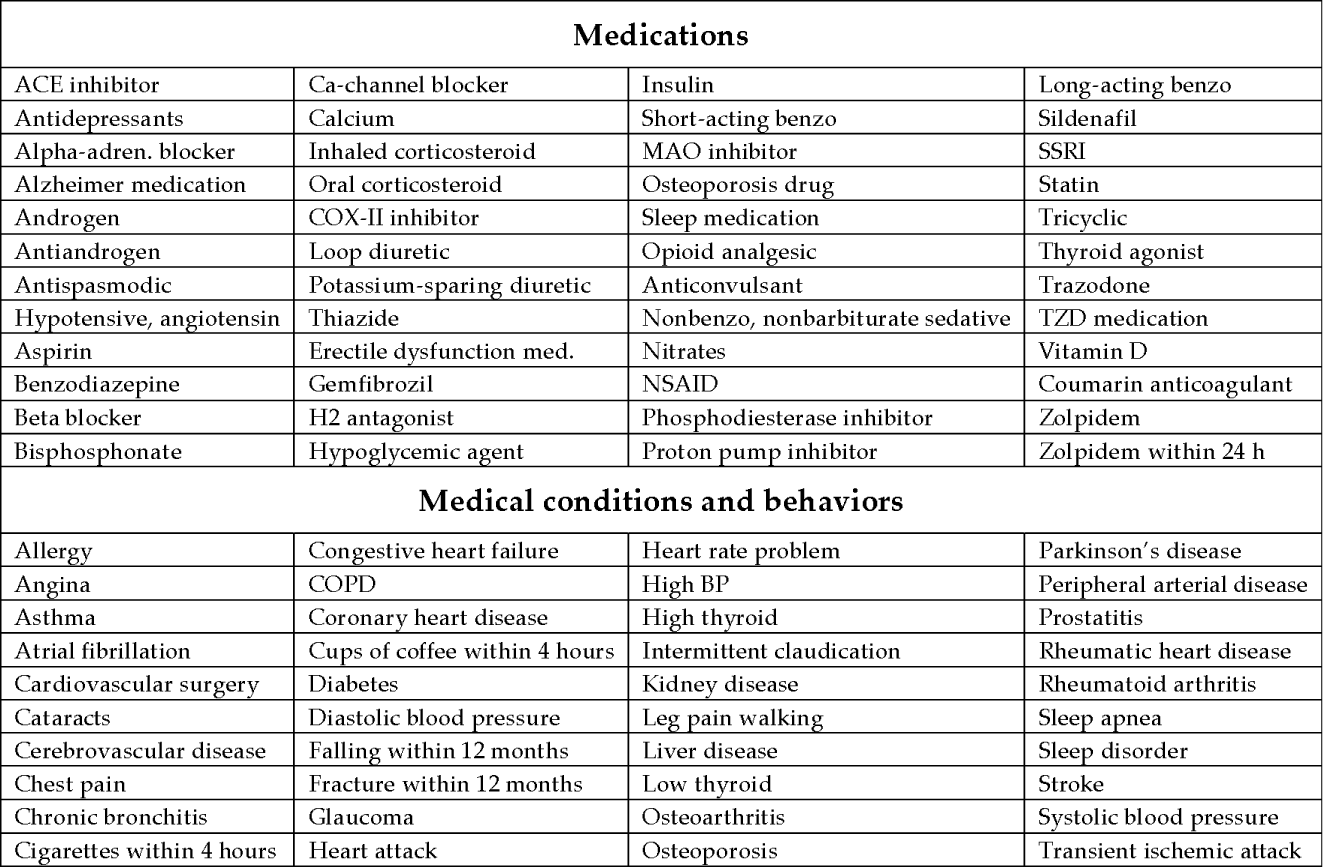
Health indicators in the analyses.

### Statistical analysis

In the first step of analyses, we calculated partial Pearson correlations between each health outcome and each sleep macrostructure variable or relative PSD at each frequency bin and on each channel, controlling for age. Point-biserial correlations were calculated in cases of dichotomous health variables. Age was controlled because changes in PSD and a worsening of health are expected as a function of aging, rendering age a strong potential confounder of any health-PSD correlation. All correlation coefficients were corrected for false detection rate (FDR) using the Benjamini-Hochberg algorithm [29]. FDR correction was applied across the 15 macrostructure variables in case of macrostructure correlations and across frequency bins for PSD correlations.

In the next steps, performed only for PSD, we aimed to identify patterns in the correlations between health and PSD. In order to accomplish this, we performed principal component analysis (PCA) on the correlation coefficients obtained in the first step for PSD variables, considering frequency bins (reduced to 48 bins by averaging to simplify analyses) as indicator variables and health indicators as observations. Correlation coefficients from recording locations C3 and C4 were averaged before PCA, while NREM and REM were treated as separate analyses. Principal components were retained based on Kaiser’s rule (eigenvalue>1). This step identified typical changes in the spectral composition of the sleep EEG when comparing those with certain health indicators to those without them.

In the final step, we extracted the principal component scores of individual health indicators on each of the extracted PCs. Health indicators were cluster analyzed based on NREM and REM PCA scores and the number of significant correlations (five variables in total) using a K-means algorithm. This step highlighted to what extent individual health indicators conform to one or more of the stereotypical patterns of PSD change identified by PCA, and if they can be grouped into meaningful clusters based on the similarity of their PSD correlation patterns.

All analyses were performed in MATLAB 2022a. Supplementary data is available at https://zenodo.org/records/10118960. Raw data is available from www.sleepdata.org.

## Results

### Descriptive statistics

Missingness of data was minimal, except for fractures (missing N=2143). For the other variables, 0–2.5% of data was missing.

The proportion of participants reporting medical conditions ranged from 0.6% (Parkinson’s disease) to 49.5% (high blood pressure) of the sample. The use of specific medications ranged from 0.1% (MAO inhibitors) to 62% (Vitamin D) of the sample.

Detailed descriptive statistics, including comorbidities (overlaps between each pair of binary health indicators) are reported in the **Supplementary data**.

### Macrostructure correlations

Close to a majority (43 out of 88) of health indicators were significantly associated with at least one sleep macrostructure variable. REM sleep was the most frequently impacted, with increased REM latency without wake (for 31 health indicators), REM latency (for 24 health indicators), and reduced REM duration (for 16 health indicators) being the most common findings. Furthermore, reduced sleep efficiency was found for 14 health indicators, and increased sleep latency for 11. While decreased REM latency or increased REM duration was not unequivocally found for any health indicator, sleep efficiency was increased in those taking trazodone and calcium and sleep latency was reduced in those taking calcium. Based on the sum of absolute correlations, antidepressant use, calcium supplementation, smoking before sleep, and benzodiazepine use were the health indicators most strongly associated with sleep macrostructure. Generally, all correlations were modest, with the largest correlation at r=0.269 (between SSRI use and REM latency without wake) and a mean absolute correlation 0.025 across all macrostructure variables and health indicators.

Figure 1 illustrates the relationship between sleep macrostructure and health indicators. Univariate correlations for all variables are available in the **Supplementary data**.

**Figure 1.**
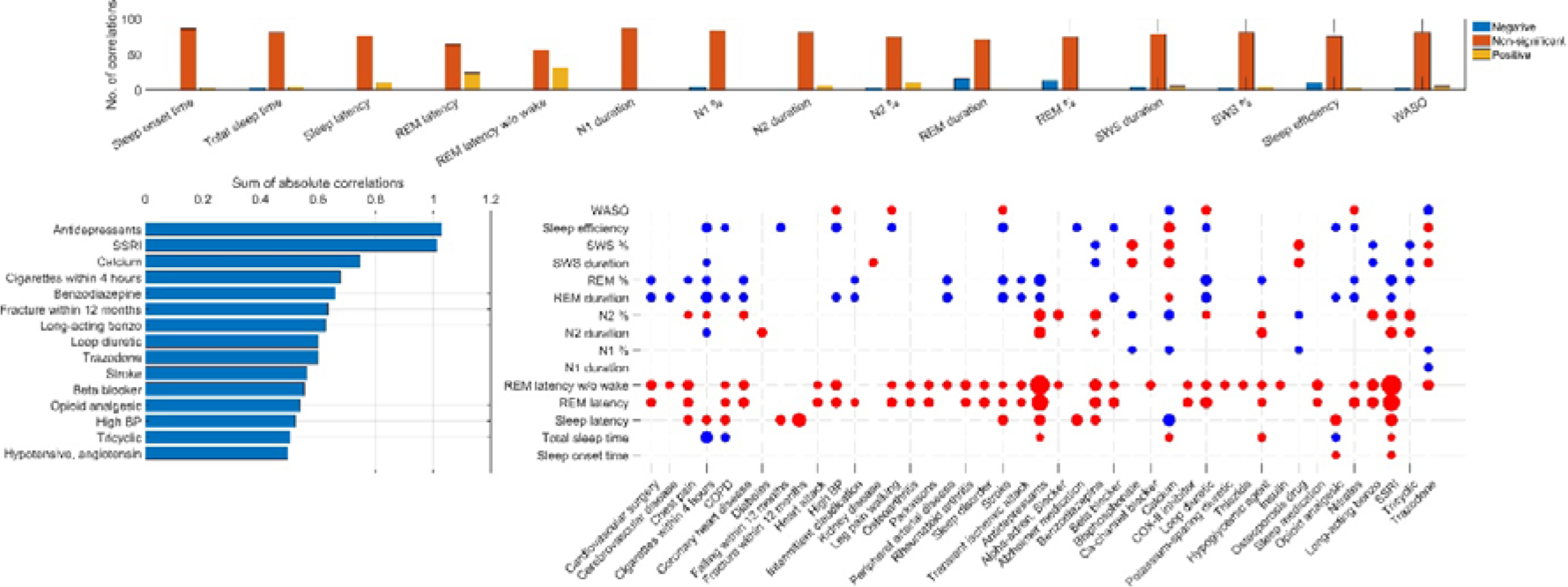
The relationship between sleep macrostructure and health indicators. **Panel A** shows the number of significant (after FDR correction) negative, non-significant and significant positive correlations with each macrostructure variable out of the 88 health indicators. **Panel B** illustrates the 15 variables with the strongest association with sleep macrostructure, expressed as the sum of absolute correlations across all macrostructure variables). **Panel C** illustrates all individual correlations between sleep macrostructure and health indicators. Significant correlations (after FDR correction) are shown with a colored circle, the radius of which is proportional to the absolute correlation value. Red circles indicate positive and blue ones negative correlations. For non-significant correlations no circle is shown, and health indicators with zero significant macrostructure correlations are not shown.

### Univariate PSD correlations

About 40% (34-42 depending on EEG recording location and vigilance state) of the 88 health indicators investigated were completely unrelated to sleep EEG PSD, as evidenced by the preponderance of zero FDR-corrected correlations (Figure 2). The frequency ranges most commonly associated with health indicators were low (<5 Hz) and intermediate (∼15-30 Hz) frequencies, with an additional substantial NREM-specific peak in the sleep spindle frequency range (∼12-15 Hz).

**Figure 2.**
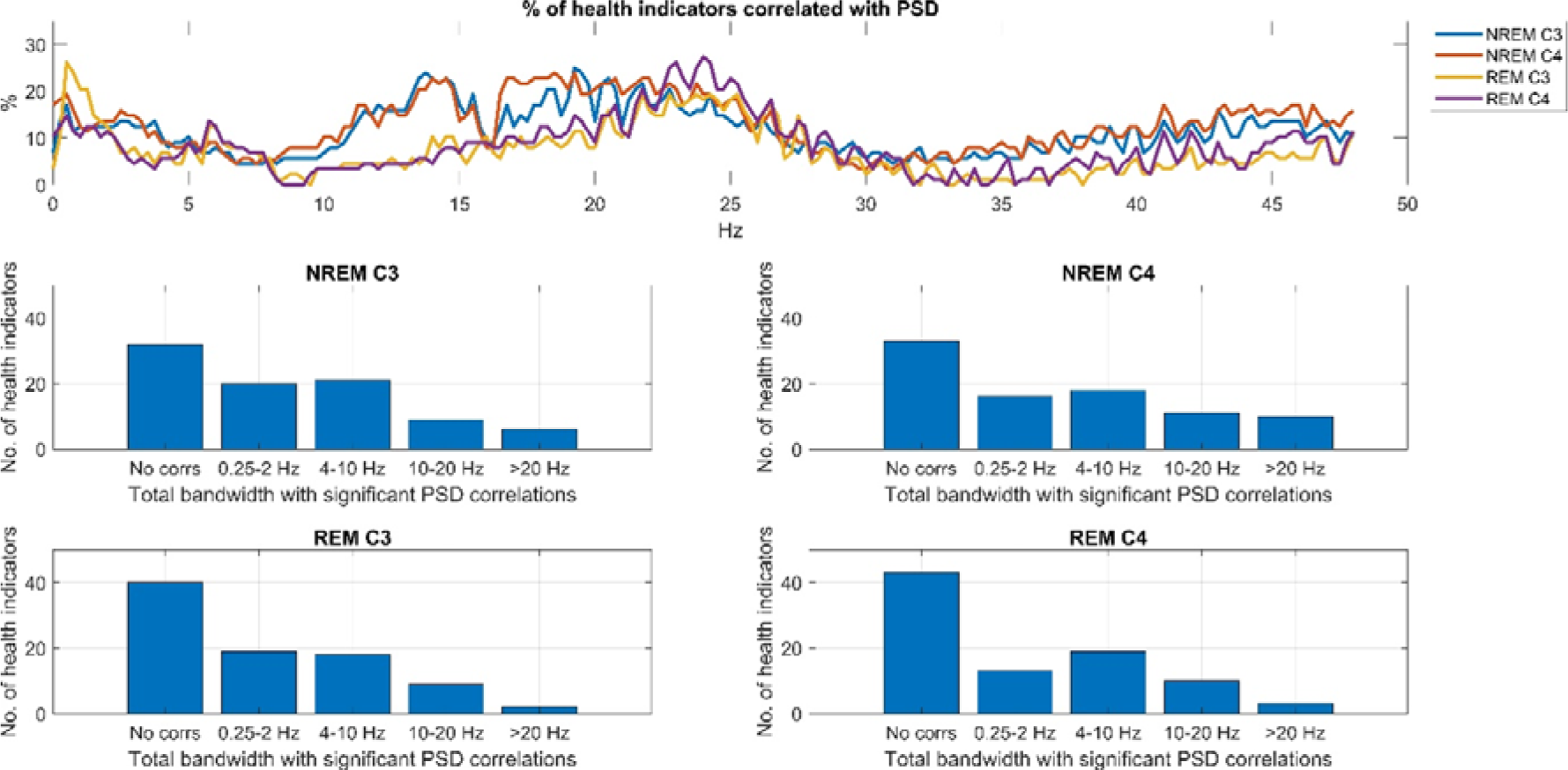
The relationship between sleep EEG PSD and health indicators. **Panel A** shows the percentage of health indicators significantly (after FDR correction) with PSD as a function of frequency, recording location and vigilance state. **Panel B** illustrates the distribution of the extent of correlations with EEG PSD. The total bandwidth refers to the total number of frequencies, regardless of continuity, exhibiting significant FDR-corrected correlations with a health indicator. Note that for a substantial number of health indicators no significant PSD correlation was seen, but a small minority of health indicators correlated with PSD in a total range of over 20 Hz.

Interestingly, health indicators with zero or close to zero associations to EEG PSD included many with presumably great effect on sleep, such as the presence of sleep disorders, sleep apnea or self-reported cigarette smoking and coffee consumption on the day before sleep (Figure 3).

**Figure 3.**
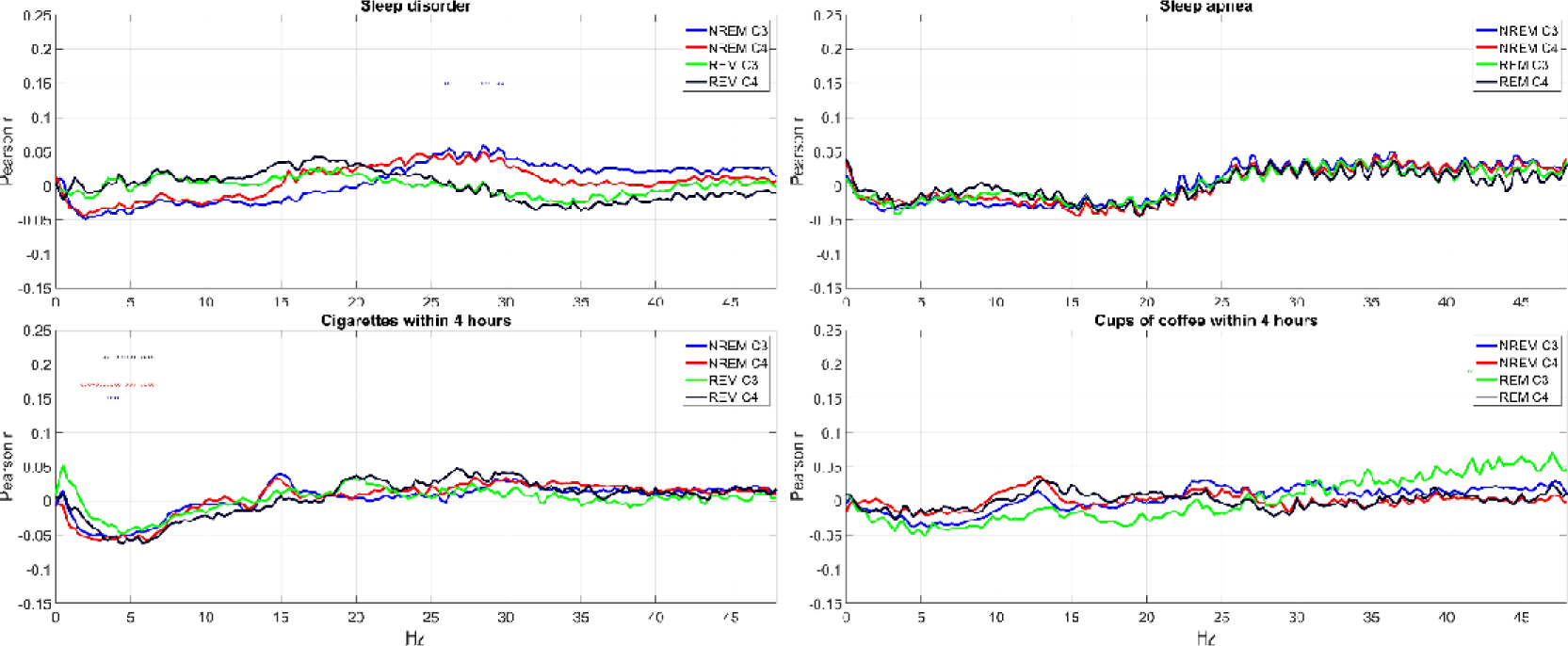
Selected health indicators with weak associations with EEG PSD. The charts show Pearson correlations (for binary variables, corresponding to point-biserial correlations) between relative log10 PSD and the self-reported presence of any sleep disorder, sleep apnea, cigarettes smoked and cups of coffee consumed within 4 hours of sleep. Correlations which are significant after correction for FDR are marked with a circle with a color marking the appropriate channel and vigilance state.

On Figure 4, we illustrate four health indicators which, in contrast to these, demonstrated a signifiant correlation with sleep EEG PSD. Medication with zolpidem was associated with increased PSD in the sigma band, corresponding to sleep spindles, selectively in NREM. Medication with selective serotonin reuptake inhibitors (SSRIs) was associated with decreased PSD in the low, and increased PSD in high frequencies. Atrial fibrillation and medication with coumarin anticoagulants were associated with an increase in the lowest and highest, and a reduction of middle frequencies. All univariate correlations are illustrated in a similar manner in the **Supplementary data**.

**Figure 4.**
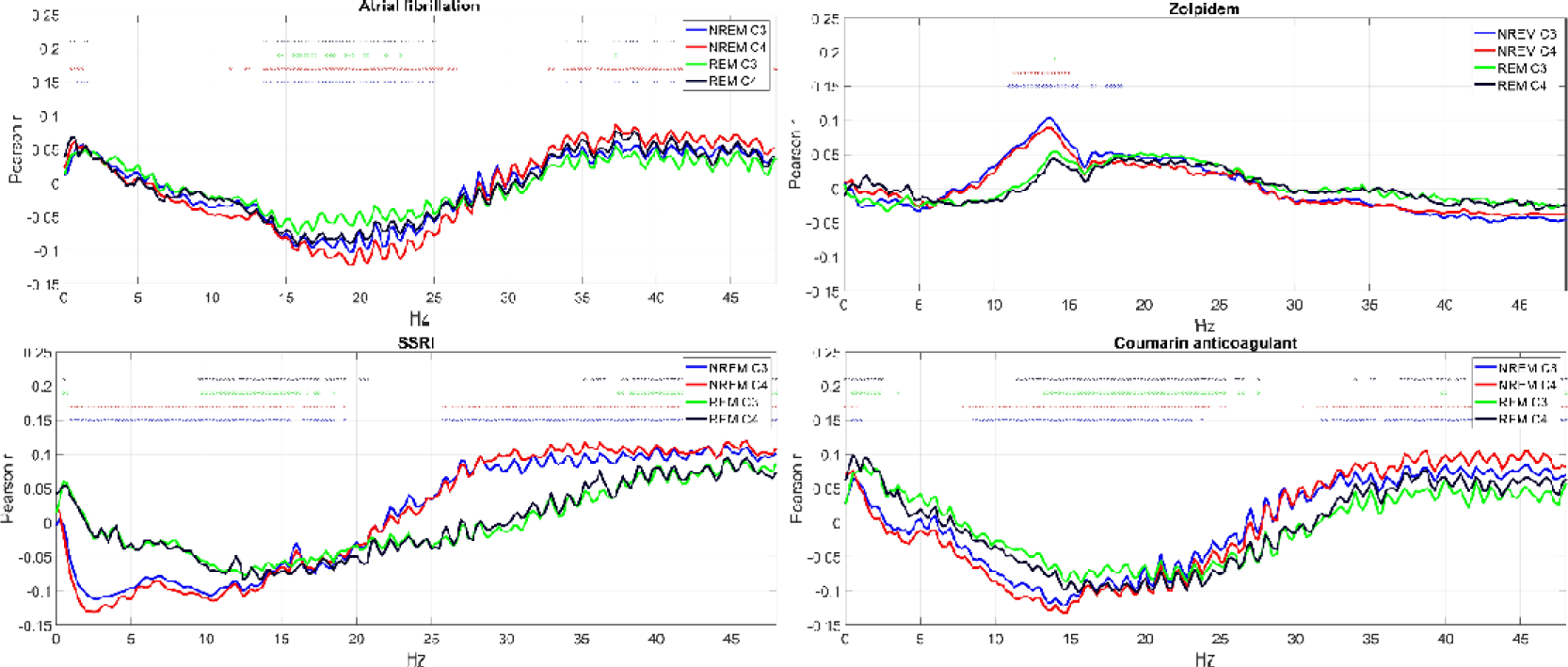
Selected health indicators with strong associations with EEG PSD. The charts show Pearson correlations (for binary variables, corresponding to point-biserial correlations) between relative log10 PSD and the self-reported diagnosis of atrial fibrillation, and medication with zolpidem, SSRIs and coumarin anticoagulants. Correlations which are significant after correction for FDR are marked with a circle with a color marking the appropriate channel and vigilance state.

In subsequent analyses, we sought to identify patterns in the non-zero PSD correlations we observed.

### Principal component analysis of PSD correlations

We found that the association of PSD in the 193 frequency bins with health indicators is highly stereotypical, with large sets of bins co-correlating with the same disorders (Figure 5, Panel A). Therefore, in the next step of our analyses, we applied PCA to the univariate health-PSD correlations we obtained to reduce the dimensionality of these variables. We found only two PCs accounted for approximately 90% of the variance. This phenomenon, as well as the loadings on the two PCs were similar across NREM and REM vigilance states. PC loadings of frequency bins on the top PCs are shown on Figure 4, Panel B.

**Figure 5.**
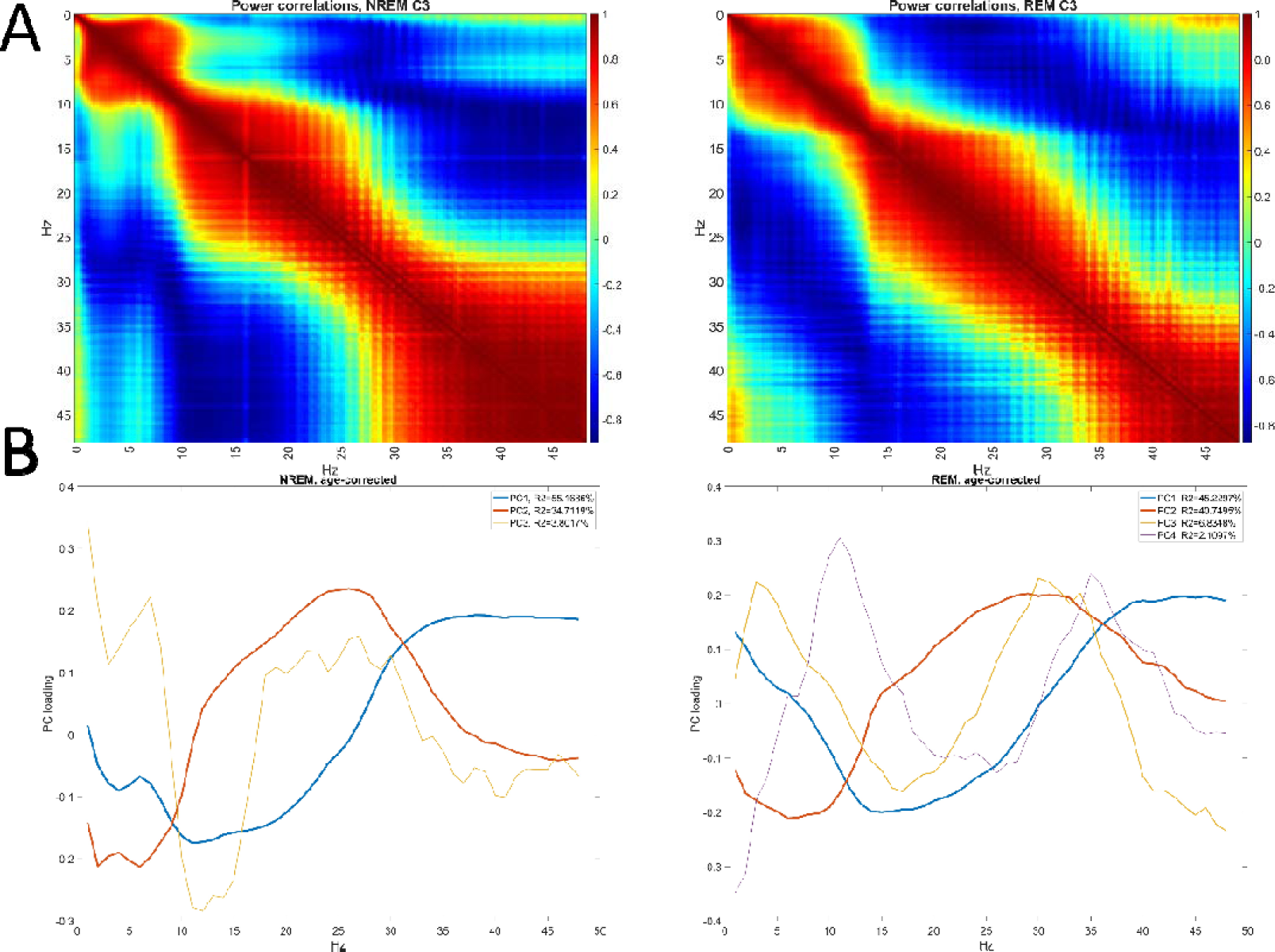
Stereotypical patterns in the health-PSD correlations. **Panel A** shows the co-correlation (Pearson correlation of correlations across health indicators) of PSD values between different frequency bins on the selected C3 channel, separately in NREM and REM. High values of this variable indicate that a pair of two PSD values tend to be correlated with health indicators in a similar manner. **Panel B** shows principal component loadings of each frequency bin in NREM and REM. Loadings are shown for all PCs with eigenvalue>1. As that the sign of loadings is arbitrary, they were multiplied by −1 in REM to optimally visualize the similarity of patterns. Line widths are proportional to the log of variance accounted for.

The strongest component had opposite-sign loadings on low (∼0-25 Hz) and high (>25 Hz) frequencies in NREM, with a similar pattern but less pronounced loadings at the lowest (∼<10 Hz) frequencies in REM. This component accounted for 55.2% of the between-bin variance in correlations with health indicators in NREM and 45.2% in REM. This component can be interpreted as an acceleration of EEG rhythms, with a loss of low and a relative increase in high frequencies, possibly signifying the loss of sleep depth or an increase in noise.

The second component had negative loadings on the lowest and highest frequencies, and positive loadings on intermediate frequencies. It accounted for 34.7% of the variance in NREM and 40.7% in REM. This component can be interpreted as a change in the proportion of sigma and beta frequency relative to other rhythms.

One more PC in NREM and two more in REM were retained following Kaiser’s rule. However, these PCs were much weaker, accounting for less than 7% of the variance. While the interpretation of REM components is less straightforward, the third NREM component had opposite loadings on sigma and low-frequency power, suggesting a specific involvement of sleep spindle activity.

### Principal component scores

We next extracted the scores of individual health indicators on the extracted PCs. Strong correlations were observed between the first two PCs extracted independently from NREM and REM data (**Table 2**), confirming the visual similarity of component loadings (Figure 5) and indicating that these largely reflect vigilance state-independent changes in the spectral composition of the EEG.

**Table 2.** Correlations between PC scores of health indicators, obtained separately from NREM and REM data. Data is shown for all PCs which were retained based on Kaiser’s rule. High correlations indicate that the degree to which health indicators conformed to the stereotypical PSD change patterns revealed by the PCs was similar in NREM and REM, suggesting vigilance state-independent effects. Note that the sign of the correlation is unimportant as the sign of PC loadings is arbitrary, and that the sign of REM loadings was flipped for visualization on **Figure 5** but not for the calculations shown in this table.

|  | NREM1 | NREM2 | NREM3 |
| --- | --- | --- | --- |
| REM1 | -0.853 | 0.53 | -0.015 |
| REM2 | -0.499 | -0.823 | 0.056 |
| REM3 | -0.002 | 0.1 | -0.444 |
| REM4 | -0.059 | 0.033 | 0.762 |

Figure 6 projects specific health indicators onto the axes specified by the first two NREM PCs and the number of significant health-PSD correlations. Because REM and NREM PC scores were highly correlated, and because the first two PCs accounted for nearly 90% of all variance in both vigilance states, these two PCs illustrate health-PSD correlation patterns with relatively little loss of information. In an attempt to create groups of health indicators with similar relations to PSD, we used a K-means cluster analysis algorithm with loadings on the first 2 NREM and REM PCs and the number of significant correlations as independent variables. After initial visual inspection of the resulting 3D scatterplot, we specified 3 clusters to be extracted. Health indicators are color coded by cluster membership. An interactive, rotatable MATLAB figure from which these figures were extracted and a video showing its rotation are available in the **Supplementary data**.

**Figure 6.**
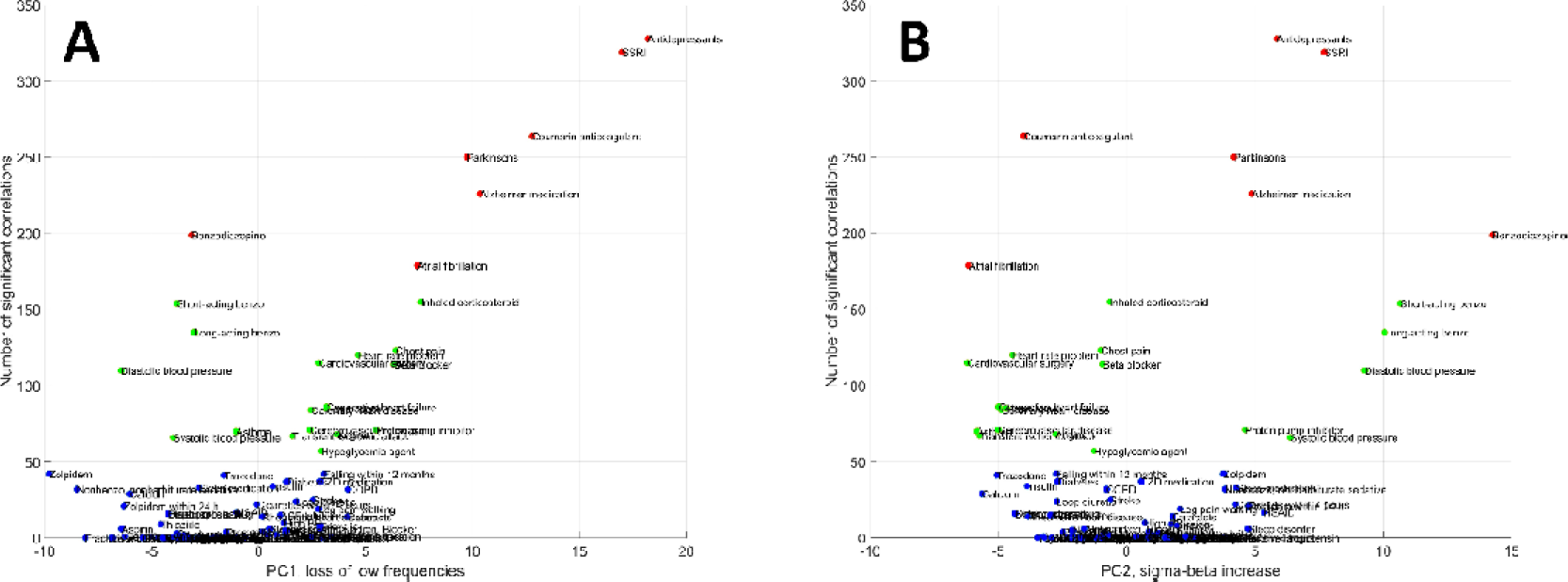
The position of individual health indicators as a function of PSD correlation PCs and the number of significant PSD correlations. The 3D scatterplot is rotated to highlight scores on specific PCs. **Panel A** illustrates scores on PC1 while **Panel B** illustrates scores on PC2. Marker colors are based on cluster membership: blue for Cluster 1, green for Cluster 2 and red for Cluster 3.

Cluster 1 contained the vast majority of health outcomes (N=63, 71.59%). Health indicators in this cluster had at most few significant correlations with PSD and low loadings on both PCs. These health indicators can be characterized as having retained sleep.

Cluster 2 was characterized by a larger number of significant health-PSD correlations and high absolute loadings on PC2. It consisted of 18 health conditions (20.45%). Interestingly, both negative and positive loadings were observed. Health indicators in Cluster 2 with negative PC loadings tended to be those associated with cardiovascular conditions: angina pectoris, cardiovascular surgery, cerebrovascular disease, chest pain, congestive heart failure, heart attack, heart rate problem, and transient ischemic attack. Atrial fibrillation exhibited a similar pattern, but was formally classified as Cluster 3 due to more correlations, while asthma and hypoglycemic agent use was also classified as Cluster 2 with negative PC2 loadings. These disorders were characterized by the specific loss of mid-frequency (sigma-beta) activity. Selected members of Cluster 2 with positive PC2 loadings are illustrated on Figure 7.

**Figure 7.**
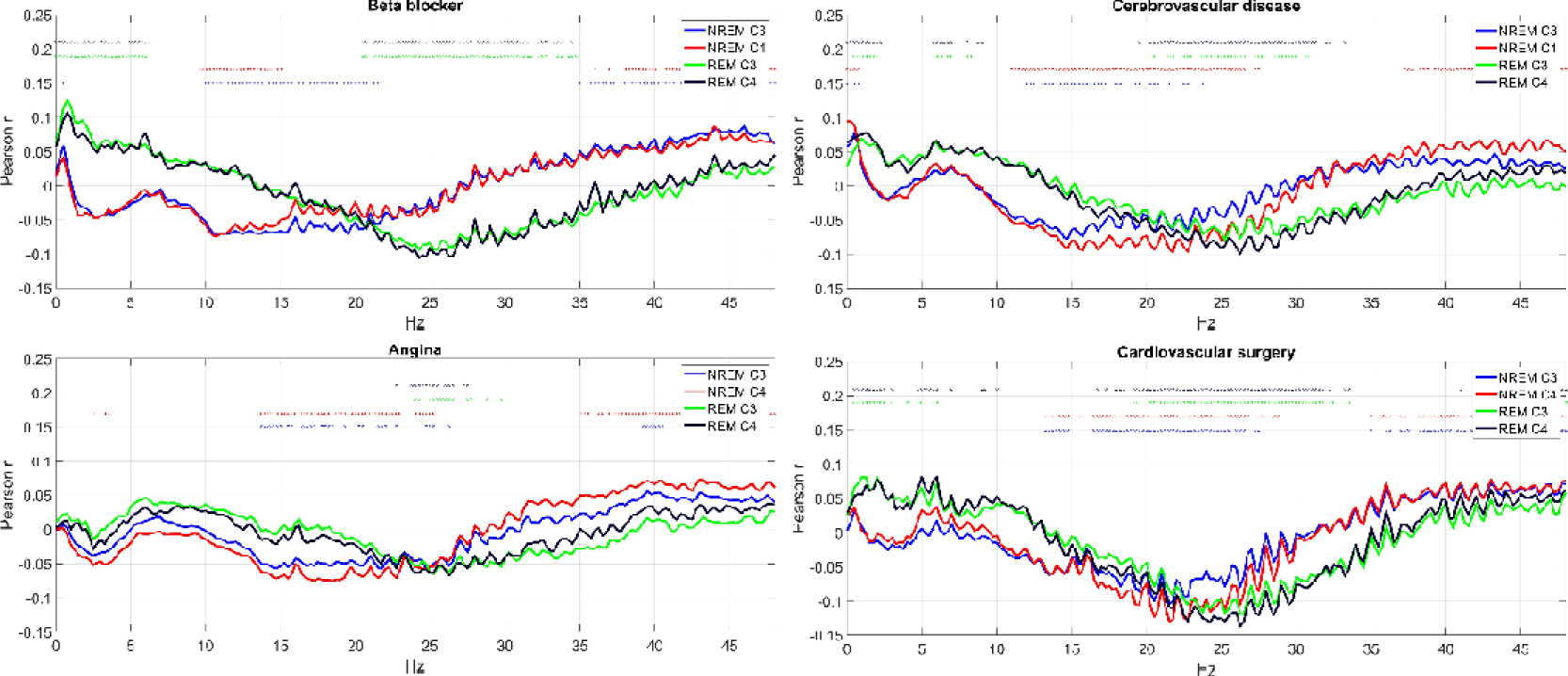
Selected health indicators in Cluster 2 with negative PC2 loadings. Axis Y shows the point-biserial correlation between health indicators and sleep EEG PSD. Correlations significant after correction for multiple comparisons are marked with a dot in the appropriate color above the lines. Note the negative correlation with mid-frequency activity.

Health indicators in Cluster 2 with positive PC2 loadings were short-acting and long-acting benzodiazepine use, systolic and diastolic blood pressure and proton pump inhibitor use. Benzodiazepine use in general exhibited a similar pattern but due to more correlations was formally classified as Cluster 3. In these conditions, the specific increase of mid-frequency (sigma-beta) activity was characteristic. Selected members of Cluster 2 with positive PC2 loadings are illustrated on Figure 8.

**Figure 8.**
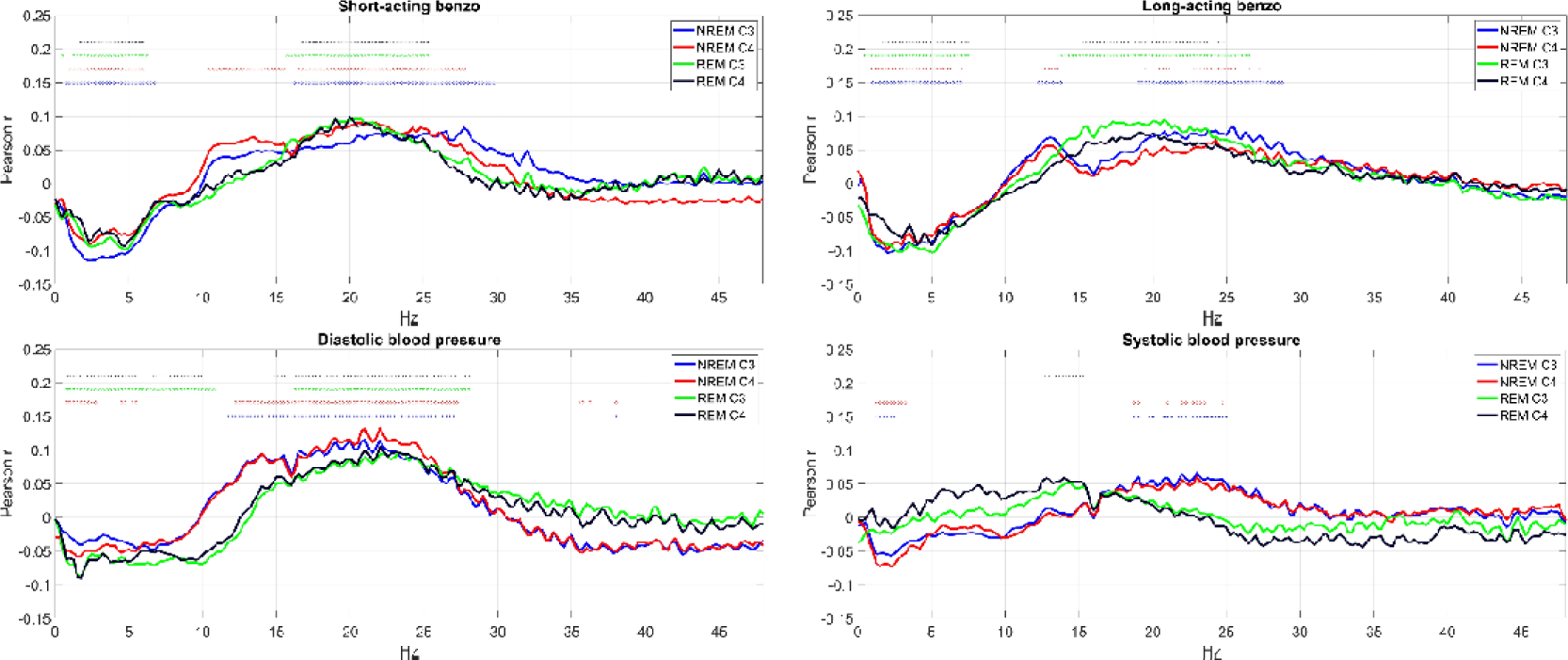
Selected health indicators in Cluster 2 with positive PC2 loadings. Axis Y shows the point-biserial correlation between health indicators and sleep EEG PSD. Correlations significant after correction for multiple comparisons are marked with a dot in the appropriate color above the lines. Note the positive correlation with mid-frequency activity.

Cluster 3 was characterized by a large number of significant health-PSD correlations and particularly high loadings on PC1. It consisted of seven health conditions (7.95%). Health indicators in this cluster were the following: antidepressant medication, SSRIs, Parkinson’s disease, Alzheimer medication, coumarin anticoagulant use, atrial fibrillation and benzodiazepine use. Benzodiazepine use, however, was an atypical cluster member due to its negative PC1 loading. Due to the pattern of PSD loadings on PC1, the other health indicators in the cluster can be characterized to have accelerated sleep EEG, with fewer low and more high frequency components. Selected members of this cluster are illustrated on Figure 9.

**Figure 9.**
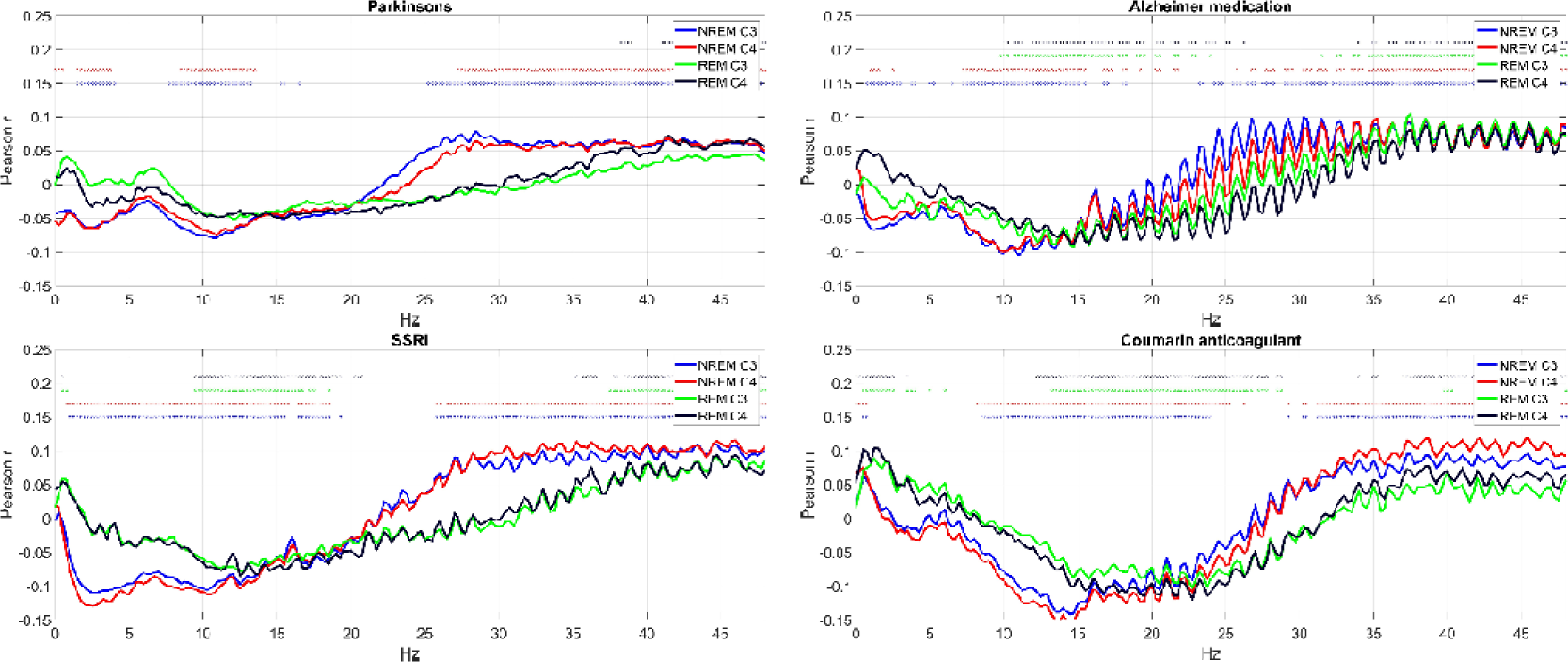
Selected health indicators in Cluster 3. Axis Y shows the point-biserial correlation between health indicators and sleep EEG PSD. Correlations significant after correction for multiple comparisons are marked with a dot in the appropriate color above the lines. Note the negative correlation with low-intermediate and the positive correlation with high frequencies.

### NREM sigma alterations in sleep medication users

We previously used PCA and cluster analysis to reduce the dimensionality of our data and find patterns in the association between health indicators and EEG PSD. While our findings confirmed that these associations are stereotypical and provided evidence that meaningfully distinct groups of health conditions with characteristic relationships to sleep exist, they did not capture all details of the association between sleep and health. Specifically, strong correlations between specific indicators and specific EEG frequencies may have been missed by these techniques as PCA identifies broad-frequency patterns which co-occur across many health indicators and cluster analysis also relies on the number of correlations between health indicators and PSD. Upon reviewing univariate associations, we discovered that sleep medications (benzodiazepines and zolpidem) are specifically associated with increased activity in NREM sigma frequencies, most likely indicating sleep spindles.

Three variables (“Zolpidem”, “Zolpidem within 24 hours” and “Nonbenzo, nonbarbiturate sedative”) indicated zolpidem use. These variables were strongly related, with 97% of those taking nonbenzo, nonbarbiturate sedatives also reporting regularly taking zolpidem and 68% of those taking zolpidem also taking it within 24 hours (**Supplementary data**). All three variables were classified as Cluster 1 (no substantial sleep alterations). However, they had relatively strong negative PC1 loadings and in univariate analyses all showed significant, specific positive correlations with NREM sigma-frequency activity.

Benzodiazepine use was indicated by three logically nested variables (“Short acting benzodiazepine” or “Long acting benzodiazepine” use, with “Benzodiazepine use” considered positive in both cases). While the first two variables were classified as Cluster 2 and the third as Cluster 3 due to the more widespread PSD correlations of the latter, all three had a similar pattern of association with PSD. Negative PC1 and positive PC2 loadings were observed, indicating an acceleration of EEG activity with a specific increase in the intermediate frequencies. In univariate analyses, however, it became clear that beyond a general, vigilance state-independent increase in the intermediate frequencies, a NREM-specific increase in sigma frequency activity is associated with benzodiazepine use, likely suggesting that these medications increase sleep spindling.

Figure 10 illustrates univariate associations between zolpidem use, benzodiazepine use, and PSD.

**Figure 10.**
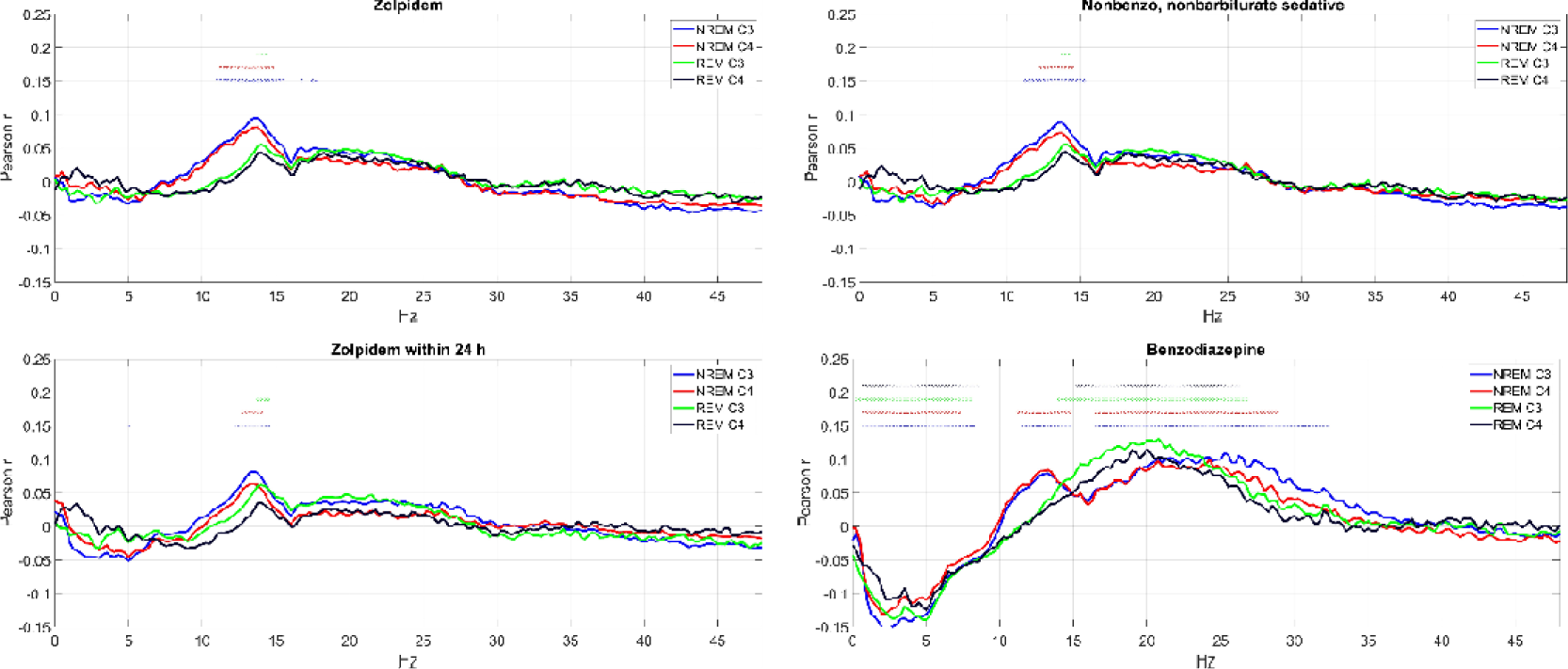
The correlation between PSD and zolpidem/benzodiazepine use. Axis Y shows the point-biserial correlation between health indicators and sleep EEG PSD. Correlations significant after correction for multiple comparisons are marked with a dot in the appropriate color above the lines. Note that a NREM-specific increase in sigma activity is associated with the use of these medications, but it accompanied by reductions in low-frequency and increases in high-frequency activity in benzodiazepine users only.

### Separating disorder and medication effects

In our final analyses, we aimed to establish whether changes in EEG PSD are associated with health problems, or the medications used to treat them. This is an issue in case of five health indicator clusters: 1) Alzheimer’s disease, 2) Parkinson’s disease, 3) depression/antidepressant use, 4) benzodiazepine use, and 5) heart disease.

In case of Alzheimer’s and Parkinson’s disease, disease and treatment effects could be separated by comparing the sleep of treated and untreated patients. However, for Alzheimer’s disease, only disease-specific medication (N=56) data is available for this wave of MrOS, rendering such an analysis unfeasible. For Parkinson’s disease (N=19), 14 participants reported taking dopaminergic medication, likely for this condition, and 5 did not. The small size of this sample also precludes definitive analyses separating medication and disorder effects.

To separate the effects of depression from antidepressant use, we calculated correlations between scores on the Geriatric Depression Scale, also available in MrOS, in participants not reporting antidepressant use. Even in antidepressant-naïve participants, increased REM latency, reduced REM duration, and an acceleration of EEG rhythms was seen, much like in SSRI users (**Supplementary figure S1**), suggesting disease as opposed to a drug effects. However, some effects (increases in WASO and reductions in sleep efficiency, a relative increase in alpha-frequency EEG rhythms) were only seen in relation to GDS scores, while the increase in REM latency and reduction in REM duration was more prominent in relation to antidepressant use.

Only SSRI use was associated with characteristics PSD changes, the use of other antidepressant drugs (MAOIs, tricyclic antidepressants and trazodone) was not (**Supplementary data**, see also **Supplementary table S1** for a tabulation of antidepressant use). Additional analyses confirmed either FDR-corrected significant differences (trazodone) or trends which do not survive FDR correction (TCAs) between EEG PSD in SSRI users and the users of other antidepressants (**Supplementary figure S2**). Thus, while depressive symptoms were alone associated with reduced slow and increased high-frequency activity in the sleep EEG, this effect was especially prevalent in SSRI users, either due to a causal effect of these drugs or because of systematic differences in the characteristics of patients taking different types of antidepressants.

A fourth health indicator associated with sleep EEG PSD was benzodiazepine use. For this indicator, we had strong hypotheses about drug, rather than disorder effects. This is because benzodiazepine use was associated with similar EEG patterns previously reported in experimental studies [15,17], with reduced slow and increased fast-frequency activity and enhanced spindling. For an empirical analysis, we took advantage of the fact that benzodiazepines are not exclusively prescribed to treat sleep disorders. Of the three sleep quality scales available in MrOS, benzodiazepine users reported worse sleep on the Pittsburgh Sleep Quality Index (B=3.82, p=10 ^-41^), better sleep on the Functional Outcomes of Sleep Questionnaire (B=-0.53, p=10 ^-4^) and a trend to better sleep on the Epworth Sleepiness Scale (B=-0.63, p=0.052). (All models corrected for age.) Of the 134 participants using benzodiazepines, 34 reported a diagnosed sleep disorder but 100 did not. We hypothesized that if it is not the medication that causes sleep alterations, but the sleep disorders for which medications are taken, then significant differences will be observed between benzodiazepine users with and without sleep disorders. (While some participants may be taking benzodiazepines for sleep problems without a formal diagnosis, the cases with formal diagnoses can be assumed to be the more severe cases.) Therefore, we compared sleep EEG PSD in benzodiazepine users with or without sleep disorders in age-corrected linear models. We found no significant differences even if no correction for false discovery rate was applied (**Supplementary figure S3**), suggesting that we have indeed found not disorder, but drug effects.

Finally, sleep alterations were observed in several diseases of the cardiovascular system, mainly characterized by vigilance state-independent reductions in beta-frequency activity. As many disorders of the cardiovascular system (angina pectoris, cerebrovascular disease, chest pain, congestive heart failure, heart attack, heart rate problem, TIA) and various treatments (cardiovascular surgery, beta blocker use) were implicated, the most likely implication is that sleep alterations are the consequences of cardiovascular disorders themselves, and not the highly variable interventions used to treat them.

## Discussion

Our work aimed to replicate and extend the previous, largely experimental, literature on health-related sleep alterations in a naturalistic multivariate study of a large sample of elderly American men. By far the most numerous group of health indicators are those characterized by retained sleep EEG. This is shown by zero or very few correlations between these indicators and PSD. Surprisingly, some of these indicators are either directly related to sleep (for example, the presence of sleep disorders), or are strongly hypothesized (for example, alcohol consumption [30]) to affect it. However, statistically significant differences in the sleep EEG are still observed in nearly a majority of health indicators. Among macrostructure markers, increased REM latency and REM latency, while among EEG markers, an acceleration of EEG rhythms and specific changes in mid-frequency activity were the strongest indicators of health. We identified four groups of health conditions in which sleep changes were especially pronounced: 1) age-related disorders (Parkinson’s and Alzheimer’s disease), 2) disorders of the cardiovascular system, 3) depression and antidepressant use and 4) hypnotic use.

In line with previous reports based on small samples of patients with Parkinson’s [9] or Alzheimer’s [31] disease, we found that age-corrected sleep alterations in age-related disorders is similar to changes related to normal aging. Both disorders were characterized by an acceleration of EEG rhythms, with a specific increase in REM latency in Parkinson’s disease. Despite purported changes in cholinergic neuronal transmission causing reductions in REM sleep [31], we found that Alzheimer’s medication use was associate with reduced sleep efficiency and increased sleep latency, but not with substantial REM sleep changes.

Cardiovascular conditions were the medical diagnoses most frequently associated with sleep alterations. The typical pattern seen in these disorders was the increase in REM sleep latency, a reduction in REM duration/percentage, as well as an alteration of mid-frequency (sigma-beta) sleep EEG activity during both NREM and REM sleep. Both REM sleep [32] and mid-frequency EEG activity in rats [33] or humans [34,35] undergo substantial circadian modulation, as opposed to NREM sleep and low-frequency activity where homeostatic modulation prevails [36]. Recently, it has been suggested that immune-mediated denervation of the pineal gland underlies circadian demodulation and consequent sleep disturbance in cardiac disease patients [12]. Our finding that sleep markers under strong circadian regulation are those which are the most affected in cardiovascular disease coheres with the concept of circadian demodulation of sleep in these patients [12]. In case of adrenergic beta_1_ receptor-blockers, their known interference with pineal melatonin production [37] may be another mechanism underlying circadian demodulation and the resulting changes in REM sleep and mid-frequency EEG activity.

Our findings showed accelerated EEG rhythms, increased REM sleep latency, reduced REM sleep duration/percentage and increased N2 sleep in participants reporting ongoing treatment with SSRIs. SSRI antidepressants are known for their inhibition of the reuptake of serotonin from the synaptic cleft, thus potentiating the effect of the transmitter on pre- and postsynaptic receptors. Our findings on the overall acceleration of sleep EEG frequencies in patients treated with SSRIs confirm the overall excitatory profile of these drugs [38] and cohere with former studies performed on a low number of healthy volunteers [39–41]. We found that these effects were specific to SSRI treatment, with different patterns emerging for other antidepressants. Tricyclic antidepressants (TCAs) are known for their sedative side effects due to antagonizing muscarinic and histamine receptors, resulting in a different sleep-wake promoting effect. Trazodone exhibits antagonistic effects on 5-HT2A (serotonergic) receptors, resulting in a wake-inhibiting and slow wave sleep-promoting effect [42] and a sedative, sleep-inducing profile of this mediation [43]. TCAs and trazodone were not significantly associated with sleep EEG changes, but the former was associated with increased N2 and decreased SWS sleep and the latter was associated with increased sleep efficiency and SWS and decreased N1 sleep, in line with clinical experience. A direct comparison of patients undergoing different antidepressive treatment regimens supported that the acceleration of EEG rhythms is specific to SSRIs. However, the fact that the severity of depressive symptoms correlated with sleep alterations in a similar manner as SSRIs even in antidepressant-naïve participants suggests that the separation of the effects of SSRI treatment from that of the underlying illness is a complex issue in an ecologically valid observational study such as ours.

Finally, we found that sleep medication use in a naturalistic setting was associated with sleep changes in the way suggested by experimental studies. Both the use of benzodiazepines and zolpidem was associated with increased sleep spindle-frequency activity in NREM, suggesting a common effect of these positive allosteric modulators of the GABA_A_ receptor complex. The imidazopyridine zolpidem, with higher affinity for the α_1_ subunit of the GABA_A_ receptor complex [44], had no significant association with other frequency components. Benzodiazepine use, however, was associated with a more complex and likely detrimental pattern of sleep changes: attenuated delta and increased beta EEG activity. This is a well-known peculiarity of benzodiazepine hypnotics, termed as the EEG fingerprint of diazepam and revealed to be pharmacologically independent of its hypnogenic effects [45].

While some studies also reported the attenuation of low frequency NREM sleep EEG activity by zolpidem [16,46–48], most of these were performed on young healthy volunteers, and the low-frequency attenuation effect of zolpidem may be age-dependent [49,50]. Unlike benzodiazepines which suppressed SWS and enhanced N2, zolpidem treatment was not associated with a significant alteration of sleep structure of our subjects. Thus, the current findings suggest that the physiological characteristics of sleep are more preserved in elderly patients treated with zolpidem as compared to subjects treated with short- and long-acting benzodiazepines, whereas the enhancement of spindle frequency activity is a shared pharmaco-EEG effect of these drugs.

Our work has some limitations. First, as with all observational studies, ours has favorable statistical power but a limited ability to infer causality. This has been only partly remedied by the methods we used (e.g. establishing symptom severity effects in antidepressant-naïve participants) and we cannot always fully answer whether health-related sleep alterations are the consequence of illness or medication. Second, our study was performed in a sample of elderly American men and our findings may not automatically generalize to other populations. Third, we emphasize that our method of using K-means cluster analysis to establish health indicator clusters based on EEG PSD alterations is exploratory. While the use of this method in our view revealed well-defined and conceptually useful clusters of health indicators characterized by similar EEG PSD changes, our results are not meant to suggest that a fixed number of health indicator groups with no overlap exist.

In sum, in a multivariate analysis of a large, ecologically valid dataset we found that sleep is preserved in many disorders and pharmacological regimes. In the cases where sleep was associated with health indicators, previous experimental studies on pharmacological effects on sleep were well-replicated. While medication use itself, rather than the presence of the sleep disorders they are used to treat, likely accounts for altered sleep in hypnotic users, age-related disorders, depression and cardiovascular disorders are likely themselves causal for sleep changes. In the latter case, our findings support a recently proposed model of immune-mediated circadian demodulation.

Given the importance of well-preserved sleep in the maintenance of a good quality of life, wellbeing and economic productivity [1,3–5], as well as the crucial role of sleep in overall health [6], research revealing the complex associations between medical conditions and objective somnological indices is ideally suited to guide endeavors to maintain health in both typical and aging populations. Furthermore, replicating or extending findings of laboratory studies about specific sleep indicators affected by health in naturalistic settings can foster basic and clinical sleep research. Our work both reveals a complex picture of the relationship between sleep and health in the elderly, it replicate and extends the extant pharmaco-EEG literature in an ecologically valid setting. The hypothesized immune-mediated circadian origin of the association between reduced mid-frequency EEG activity in heart conditions suggests a need for additional studies replicating and investigating this phenomenon in depth.

## Data Availability

Raw data is available from www.sleepdata.org.

https://www.zenodo.org/records/10118960

## Acknowledgements

This research has been implemented with the support provided by the Ministry of Innovation and Technology of Hungary from the National Research, Development and Innovation Fund, financed under the TKP2021-EGA-25 funding scheme. Péter P. Ujma was supported by the National Research, Development and Innovation Office – NKFIH (grant number: 138935).

## Notes

### Competing Interest Statement

The authors have declared no competing interest.

### Author Declarations

The study used only openly available human data that were originally located at www.sleepdata.org.

